# Evaluating seasonal variations in human contact patterns and their impact on the transmission of respiratory infectious diseases

**DOI:** 10.1101/2022.02.22.22271357

**Authors:** Allisandra G. Kummer, Juanjuan Zhang, Chenyan Jiang, Maria Litvinova, Paulo C. Ventura, Marc A. Garcia, Alessandro Vespignani, Huanyu Wu, Hongjie Yu, Marco Ajelli

## Abstract

Considerable uncertainty surrounds the seasonality of respiratory infectious diseases. To which extent the observed seasonality is associated with biological reasons (e.g., virus survival rates, host immune dynamics) or human behavior remains unclear. Here, we investigate the association between temperature and human contact patterns using data collected through a contact diary-based survey between December 24, 2017, and May 30, 2018, in Shanghai, China. We identified a significant inverse relationship between the number of contacts and both the seasonal temperature trend (p=0.003) and daily temperature variation (p=0.009). The average number of contacts increased from 18.9 (95% CI: 14.5-21.6) in December to 20.9 (95% CI: 15.4-26.5) in January before decreasing to 11.6 (95% CI: 8.7-14.8) in May. This seasonal trend in the number of contacts translates into a seasonal trend in the reproduction number – the mean number of secondary cases generated by a typical infector. We developed a compartment model of influenza transmission informed by the derived seasonal trends in the number of contact patterns and validated against A(H1N1)pdm09 influenza data for the focus location and study period. We found that the model shows an excellent agreement with the estimated influenza dynamics providing support to the hypothesis that the seasonality in contact patterns shapes influenza transmission dynamics. Our findings contribute to a deeper understanding of the epidemiology of respiratory infectious diseases and could potentially inform improved preparedness planning.

## Introduction

Several respiratory infectious diseases, including influenza and respiratory syncytial virus (RSV), show clear seasonal trends and cyclic epidemics [1-3]. Specifically, temperate regions experience the highest incidence of these diseases during the winter seasons with fewer cases occurring during the summer months whereas places with tropical climates, such as Singapore, may observe a higher incidence of disease in warmer months or year-round [1, 3-7]. Possible explanations for the seasonality of respiratory infectious diseases include the variations in meteorological conditions (e.g., absolute humidity, temperature) that influence virus transmission, survival, and host susceptibility [5, 8, 9]. Previous studies have indicated that influenza virus survival is associated with water vapor in the air, suggesting that absolute humidity (a measurement of water vapor regardless of temperature) might be a driver of the seasonality of influenza epidemics seasonality [8, 9]. This finding is supported by some epidemiological studies [8-10], but not by others [11-13], suggesting that this may be a contributing factor but not the main driver of influenza seasonal behavior. For instance, seasonal changes have also been linked to changes in host immune function, particularly decreases in mucosal integrity during dry seasons may increase susceptibility to infection [4, 6]. Overall, the underlying mechanism(s) that drive seasonality remain elusive [14].

A large amount of work has been done identifying human contact patterns as a key determinant for infectious disease transmission [5, 15-18]. For example, contacts in locations such as schools and workplaces tend to have the highest rates of transmission due to close contacts [5, 19-21]. While contacts made in schools and workplaces are relatively consistent, human behavior adapts to contextual changes due to working days, weekend days, holidays, and weather conditions [5]. Seasonal trends in human behavior could then be one of the drivers underlying the seasonality of disease transmission [14]. School openings and closures during holiday breaks reduce the number of contacts among school-age children which may explain why influenza is reduced during holiday breaks [13, 15, 16, 19]. Furthermore, people tend to spend more time indoors when the temperature drops [5, 22], increasing an individual’s proximity to others thus impacting their likelihood of contracting an infectious disease [23, 24].

In this work, we investigate whether human contact patterns follow seasonal trends and the extent to which these trends can contribute to shaping the seasonal patterns observed for respiratory infectious diseases. To this aim, we collected contact survey data, meteorological data, influenza-like illness (ILI) data, and A(H1N1)pdm09 influenza positivity rates for Shanghai, China, before the COVID-19 pandemic. We conducted a regression analysis showing significant associations between contact patterns and both the temperature seasonal trend and daily variations. We then leveraged the obtained results to calibrate a mathematical model of influenza transmission, which is validated against the collected influenza data for Shanghai. The performed simulations ultimately provide a mechanistic explanation of the observed influenza seasonality based on the seasonality of human contact patterns.

## Results

### Contact Patterns

Analyses were conducted to investigate the seasonality of human contact patterns for 965 participants using data collected through a diary-based contact survey [25] and daily temperatures in Shanghai between December 24, 2017 and May 30, 2018. A total of 18,116 close contacts (mean = 18.7 per day per participant, Interquartile Range (IQR): 4.0 to 30.0) were analyzed (Table 1). The number of participants per week ranged from 0 to 157, and the total number of contacts each week was in the 0-3,331 range (Fig. S1). Seasonality was represented through temperature seasonal trends and daily temperature variation. The seasonal trend was defined as a spline interpolating daily (maximum) temperatures, and daily temperature variation was defined as the difference between daily (maximum) temperature and seasonal trend (Fig. 1*A*). Details are provided in the Methods section and the Supplementary Material.

**Table 1.**
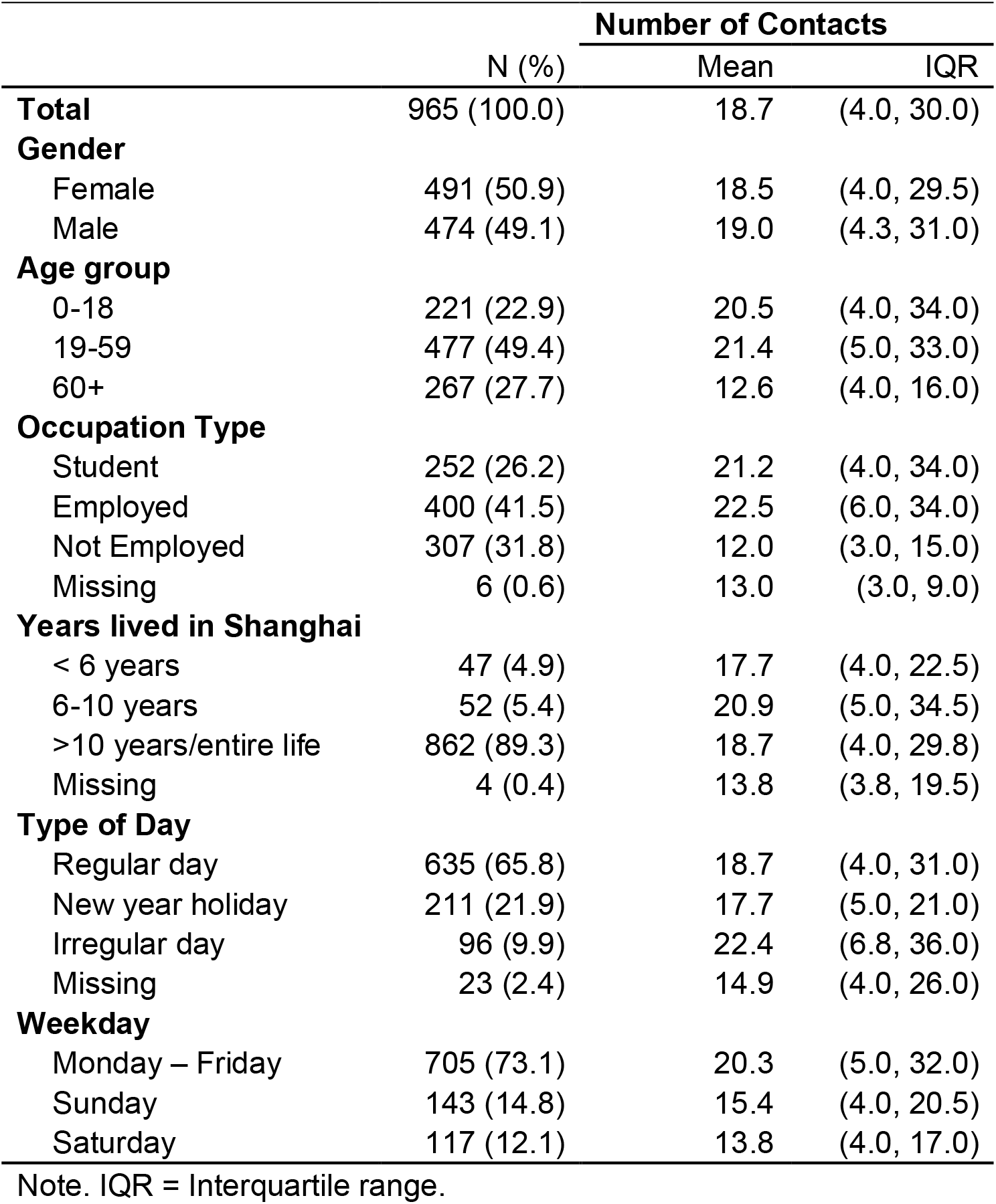
Descriptive statistics of the participants and their total contacts.

**Figure 1.**
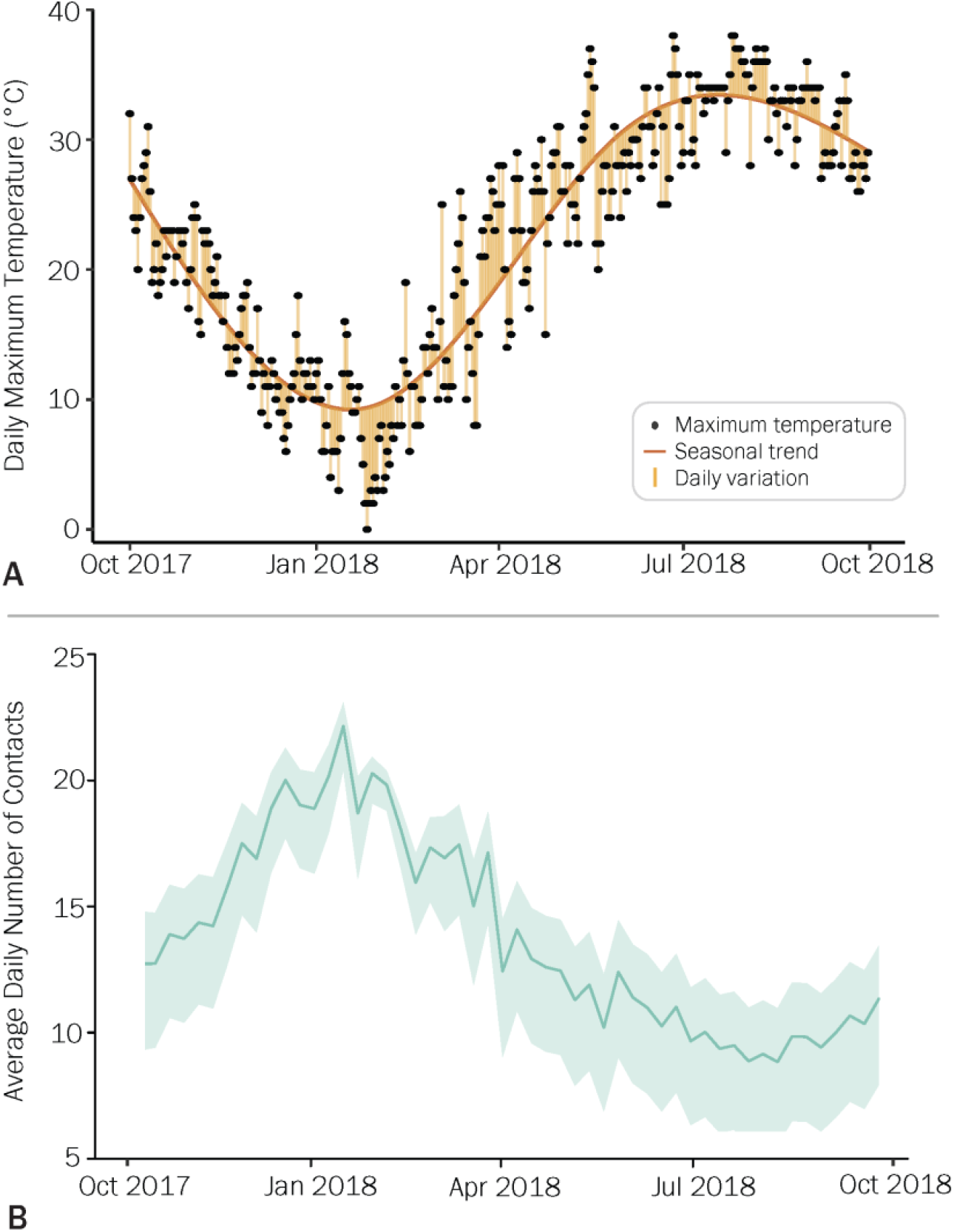
**A**. Daily maximum temperature (°C) for each day from October 1, 2017, to September 30, 2018, the seasonal trend of the temperatures, daily variation between the maximum temperature, and seasonal trend. **B**. Estimated daily number of total contacts for each week from October 1, 2017, to September 30, 2018. This estimate does not consider the potential effect of summer vacation on contact patterns. The line and shaded area represent the mean and 95% CI of the mean daily values for each season, respectively.

Age, gender, household size, occupation type, number of years lived in Shanghai, day of the week (i.e., weekday, Saturday, or Sunday), and type of day (e.g., regular day, vacation) when the contact diary was completed were included in the analysis to adjust for confounding factors. Descriptive statistics for the covariates are shown in Table 1. The study sample included slightly more female (50.9%) than male (49.1%) participants. Nearly half of the participants were between the ages of 19 and 59 (49.4%) and were employed (41.5%), while a majority had lived in Shanghai for more than 10 years or their entire life (89.3%). Adults 60 years and older reported 12.6 contacts on average (IQR: 4.0-16.0) whereas those 19-59 years had 21.4 contacts (IQR: 5.0-33.0) and participants 0-18 years old had an average of 20.5 contacts (IQR: 4.0-34.0). Employed persons reported a number of contacts similar to that of students (22.5, IQR: 6.0-34.0, vs. 21.2, IQR: 4.0-34.0, on average). Approximately, 73.1% of participants completed the diaries during a weekday (Monday-Friday) and had more contacts on average (mean: 20.3, IQR: 5.0 – 32.0) compared to Saturday (mean: 13.8, IQR: 4.0-17.0) and Sunday (mean: 15.4, IQR: 4.0-20.5). Most participants (65.8%) completed their contact diaries on a regular day (i.e., a working day not during school vacations).

### Seasonality of contact patterns

By using a multivariate negative binomial regression model with a log link function (see Methods and in the Supplementary Material for details), we found that seasonal and daily variations in temperature were associated with a significant decrease in the number of contacts (Table 2). Specifically, for each degree increase in seasonal temperature trend, there is a decrease of 0.987 (p = 0.003, 95% CI: 0.978-0.996) expected contacts. Likewise, for each degree of daily variation in temperature, expected contacts decrease by 0.981 (p = 0.009, 95% CI: 0.966-0.996) contact. By incorporating year-round temperature data into the negative binomial model, we were able to predict the number of contacts beyond the time frame of the contact survey (refer to Methods for details). As a result, we estimate an average of 18.9 (95%CI: 14.5-21.6) contacts per day in December 2017 that increases to an average of 20.9 contacts (95%CI: 15.4-26.5) in January 2018 (Fig. 1*B* and Fig. S5). The number of daily contacts declines to 11.6 (95%CI: 8.7-14.8) in May 2018 and fluctuates between 8 and 11 contacts on average throughout the rest of the year (Fig 1*B*). There is a decreasing trend in the number of contacts as the daily maximum temperature increases, possibly indicating a movement from activities in proximity with others (i.e., indoor aggregations) to more distanced activities with a lower number of contacts (i.e., outdoor activities). We found consistent results in two sensitivity analyses where we did not consider the New Year holiday period and considering the summer vacation (Fig. S6).

**Table 2.**
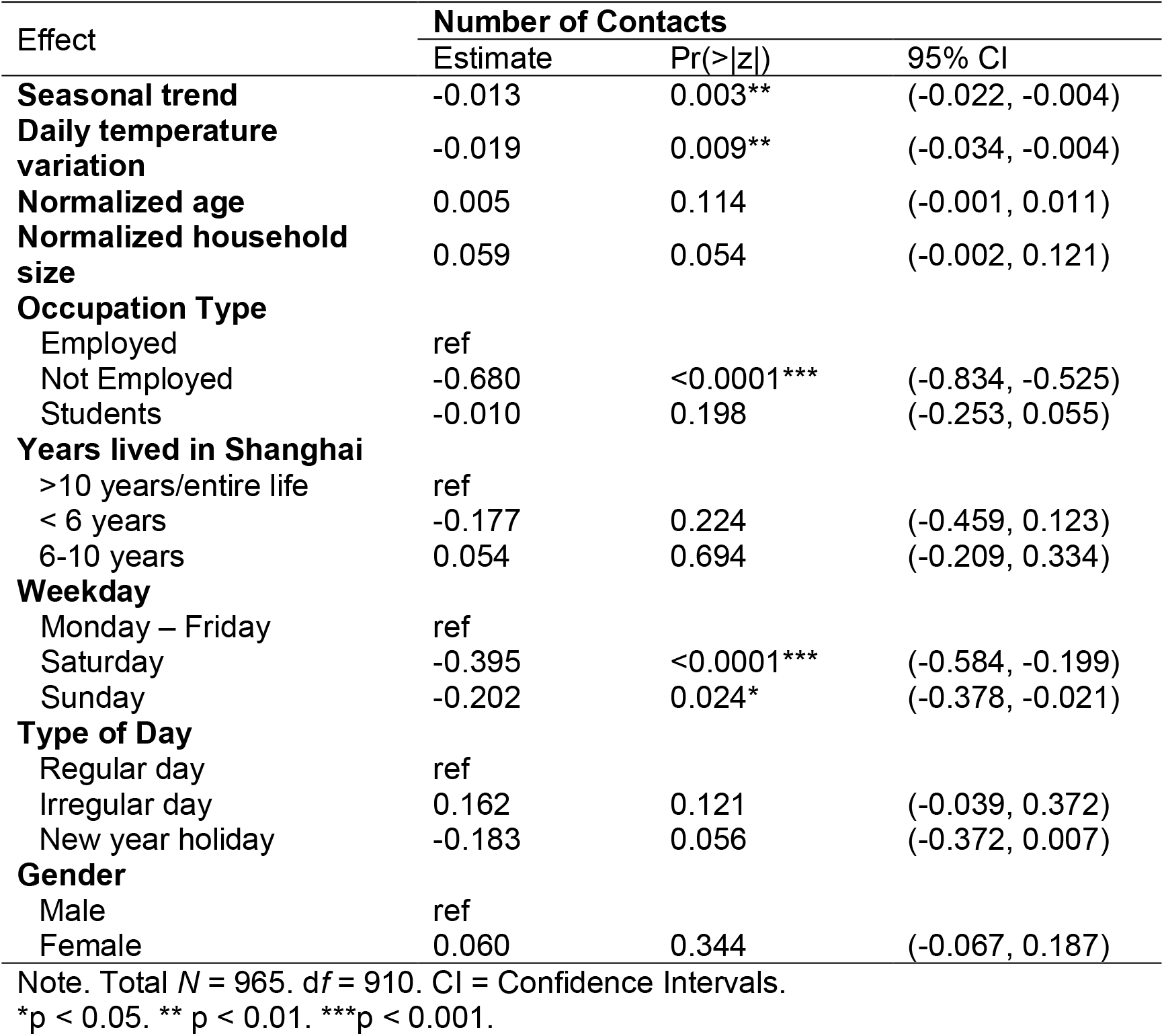
Negative binomial regression models of the effects of seasonal trend and daily temperature variation on total contacts adjusting for the covariates.

| Effect | Number of Contacts |  |  |
| --- | --- | --- | --- |
|  | Estimate | Pr(>\|z\|) | 95% CI |
| **Seasonal trend** | -0.013 | 0.003** | (-0.022, -0.004) |
| **Daily temperature variation** | -0.019 | 0.009** | (-0.034, -0.004) |
| **Normalized age** | 0.005 | 0.114 | (-0.001, 0.011) |
| **Normalized household size** | 0.059 | 0.054 | (-0.002, 0.121) |
| **Occupation Type** |  |  |  |
| Employed | ref |  |  |
| Not Employed | -0.680 | <0.0001*** | (-0.834, -0.525) |
| Students | -0.010 | 0.198 | (-0.253, 0.055) |
| **Years lived in Shanghai** |  |  |  |
| >10 years/entire life | ref |  |  |
| < 6 years | -0.177 | 0.224 | (-0.459, 0.123) |
| 6-10 years | 0.054 | 0.694 | (-0.209, 0.334) |
| **Weekday** |  |  |  |
| Monday – Friday | ref |  |  |
| Saturday | -0.395 | <0.0001*** | (-0.584, -0.199) |
| Sunday | -0.202 | 0.024* | (-0.378, -0.021) |
| **Type of Day** |  |  |  |
| Regular day | ref |  |  |
| Irregular day | 0.162 | 0.121 | (-0.039, 0.372) |
| New year holiday | -0.183 | 0.056 | (-0.372, 0.007) |
| **Gender** |  |  |  |
| Male | ref |  |  |
| Female | 0.060 | 0.344 | (-0.067, 0.187) |

Note. Total $N = 965$ . $df = 910$ . CI = Confidence Intervals.
\* $p < 0.05$ . \*\* $p < 0.01$ . \*\*\* $p < 0.001$ .

Our primary analysis utilized maximum daily temperature to gauge seasonal trends and daily variation, as individuals typically engage in activities during the warmest parts of the day. To evaluate the robustness of our findings, we conducted two sensitivity analyses employing alternative meteorological measurements: average daily temperature and absolute humidity. Trends using average daily temperature reflected those in the main analysis with the estimated monthly mean number of contacts decreasing from 24.4 (95%CI: 19.1-28.1) in January to 10.8 (95%CI: 9.9-11.7) in May (Fig. S9*B*). Results using absolute humidity showed a similar pattern, although with lower absolute values, decreasing from 22.0 contacts in January (95%CI: 17.2-24.0) to 7.0 contacts in May (95%CI: 5.7-8.3) (Fig. S9*C*).

To evaluate differences between climatic conditions, we used the estimated regression coefficients to predict the number of contacts in two alternative locations: Beijing and Guangzhou. Beijing has a humid, continental climate where summers are hot and humid and winters are cold, while Guangzhou has a humid, subtropical climate. Both locations follow similar trends as Shanghai; however, the number of contacts estimated for Beijing had a larger range between the winter and summer contacts compared to the other locations (Fig. S10 *B*). The contact patterns for Guangzhou remained relatively consistent throughout the year compared to the other locations (Fig. S10*C*).

### Seasonality of influenza

Among the different respiratory infectious diseases showing clear seasonal trends, we selected influenza in Shanghai as an illustrative example. We developed a simple homogenous-mixing compartment model of influenza transmission where the contact rate is based on the estimated number of daily contacts. The generation time was set at 3.0 days [26], while Markov chain Monte Carlo (MCMC) was used to estimate the posterior distribution of the per-contact transmission risk (β), reporting rates, and number of infected individuals at the beginning of the season by exploring the likelihood of observing the weekly number of ILI^+^ (i.e., weekly number if ILI cases multiplied by the proportion of samples positive for influenza A(H1N1)pdm09) during the 2017-2018 influenza season in Shanghai (see Methods).

The estimated mean number of ILI^+^ shows a good agreement with the reported data (Fig. 2*A*); the estimated ILI^+^ rapidly grew until nearly the end of January. The net reproduction number increased from 1.24 (95% CI: 1.21-1.27) in December to a peak of 1.34 (95% CI: 1.31-1.37) in January. Then, the epidemic started to enter a decline phase in early February when the net reproduction number fell below the epidemic threshold (Fig. 2*B*). The estimated median infection attack rate was 27.4% (95% CI: 23.7-30.5%) (Fig. 2*C*).

**Figure 2.**
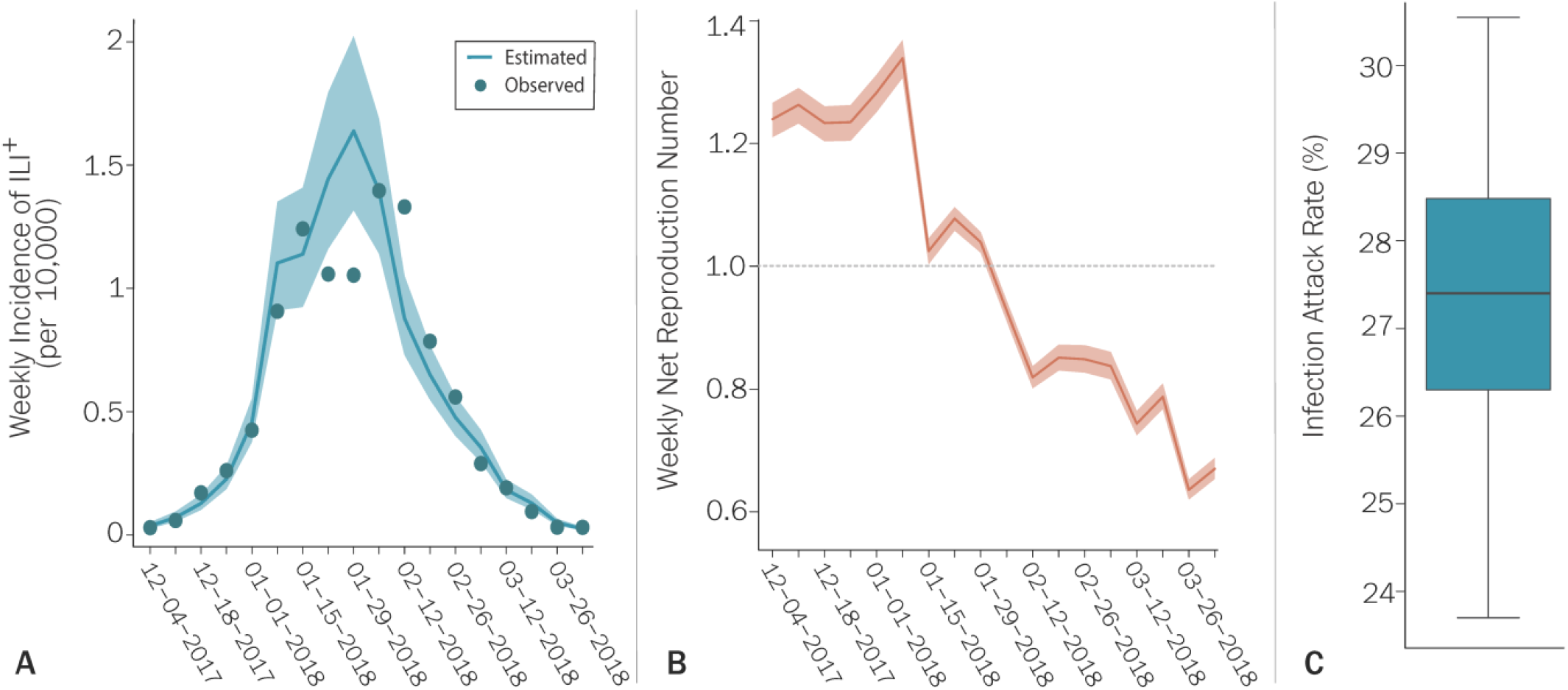
**A**. Estimated weekly incidence of ILI^+^ infections during the 2017-2018 influenza season in Shanghai, China. Dots represent the observed data; line and shaded area represent the mean and 95% CI of model simulations. **B**. Estimated posterior distribution of weekly net reproduction number. The line and shaded area represent the mean and 95% CI, respectively. **C**. Posterior distribution of the final infection attack rate for the 2017-2018 influenza season. The boxplot reports quantiles 0.025, 0.25, 0.5, 0.75, and 0.975 of the distribution.

To investigate the potential of influenza to spread in the population, we defined a new indicator: the contact-dependent basic reproduction number, defined as the number of secondary cases generated by a typical infector in a completely susceptible population at time *t*. This Indicator is equivalent to the basic reproduction number, but it considers that the number of contacts per day may be variable over time. To estimate temporal dynamics of the contact-dependent basic reproduction number, we considered the posterior distribution of the transmission risk (*β*) for the 2017-2018 influenza season and assumed the population to be fully susceptible at each time of the year. We estimated an increasing trend through the fall and winter seasons, which starts from a contact-dependent basic reproduction value of 0.81 (95%CI: 0.79-0.83) in October and increases to a peak of 1.38 (95%CI: 1.35-1.41) in January, followed by a gradual decrease through the spring season, reaching 0.71 (95%CI: 0.70-0.73) in June and remaining on average below the epidemic threshold throughout the summer (Fig. 3*A*). It is important to remark that this number is not equivalent to the effective reproductive number that considers several other factors affecting the pathogen transmissibility such as population immunity, vaccination rates, mandated or spontaneous behavioral changes in the population.

**Figure 3.**
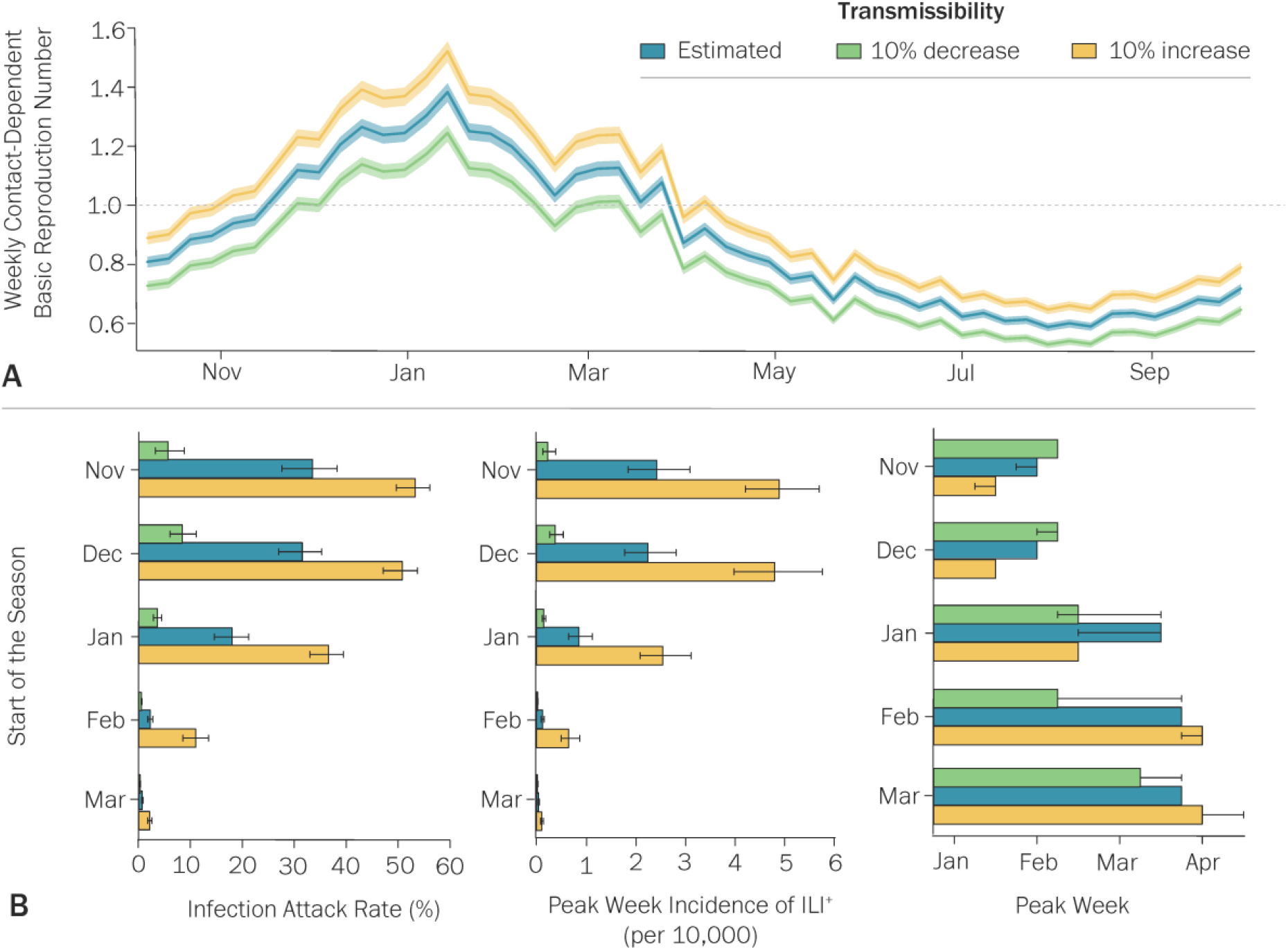
**A**. Estimated weekly contact-dependent basic reproduction number from October 1, 2017, to September 30, 2018, using the estimated posterior distribution of the transmission risk (*β*) for the 2017-2018 influenza season. The line and shaded area represent the mean and 95% CI, respectively. Scenarios considering a 10% increase and 10% decrease in transmissibility were obtained by multiplying each value sampled from the distribution of the transmission risk by 1.1 and 0.9, respectively. **B**. Infection attack rate, peak week incidence of ILI^+^, and peak week for one year of simulations for different epidemic starting times. Results are based on one thousand samples of the posterior distributions of the per-contact transmission risk and initial number of infections as estimated by MCMC for the 2017-2018 influenza season. The barplot reports mean, while the whiskers report quantiles 0.025 and 0.975 of the distribution.

To assess the effect of seasonal variations in contact patterns on influenza epidemics, we simulated a set of influenza epidemics beginning at different times of the year when the contact-dependent basic reproduction number was below the epidemic threshold (Fig 3*A*) and assuming differences in transmissibility. Namely, transmission risk was set as the estimated distribution for the 2017-2018 season, a 10% increase of transmission risk (corresponding to peak a value of the contact-dependent basic reproduction number of 1.52, 95%CI: 1.49-1.55), and a 10% decrease (peak contact-dependent basic reproduction number: 1.24, 95%CI: 1.22-1.27) (Fig. 3*B*). These scenarios are in line with the estimated values of the reproduction number of influenza, which is generally in the range 1.1-1.5 [27]. All scenarios showed the highest median infection attack rate and peak week incidence for an epidemic starting in November with the peak week in January (Fig. 3*B*).

Median infection attack rates and peak week incidence were lower as epidemics began in later months of the influenza season with peak weeks all occurring before mid-April (Fig. 3*B*).

## Discussion

The present study reveals a significant correlation between weather conditions and human contact patterns, indicating that the number of contacts is influenced by both seasonal changes and daily temperature variations. Similarly, the influenza transmission model, informed by the measured contact patterns, exhibits a pronounced seasonal trend, further highlighting this relationship as a potential contributor to the seasonality of respiratory viruses. Although no distinction is made between indoor and outdoor contact, these patterns suggest notable changes in human behavior driven by seasonal shifts. From increased proximity to others during colder weather to reduced close contacts in warmer seasons, this implies a transition from indoor to outdoor activities. This likelihood is particularly relevant for contacts occurring in community settings outside typical social environments such as homes, schools, and workplaces, due to alterations in gathering or meeting contexts. To evaluate these associations, the current study conducted an additional analysis of community contacts, which confirmed the strong connections between seasonal trends and contact patterns but not for daily temperature fluctuations (Table S3).

In the main analysis, employed persons had significantly more contacts than those who were unemployed, and working days had significantly more contacts than weekend days, indicating most contacts happen during work-related activities. Inverse associations between the number of contacts and weekend days found in the main analysis are supported by previous literature [5]. In contrast, there are significantly more community contacts on Sundays and Saturdays than during the weekend days, and unemployed persons have more community contacts than employed individuals (Table S3). Variations in weekend schedules and routines for unemployed people may allow for more opportunities to interact in the community. This may explain the significantly lower number of total contacts found for unemployed individuals.

Our analysis of the 2017-2018 influenza season in Shanghai shows a good agreement between the surveillance data and the output of the model informed with seasonal contact patterns. Furthermore, our findings about the timing of the influenza A(H1N1)pdm09 depending on the time of importation of the virus are consistent with Shanghai surveillance data for multiple seasons [28]. In addition to reproducing the temporal dynamics of influenza, the model also provides estimates of other key epidemiological indicators such as the reproduction number and infection attack rate that agree with previous estimates [27, 29-31]. These findings lend support to the hypothesis that the observed seasonality of contact patterns may play a role in driving the seasonal nature of influenza. It is crucial to recognize that our results, particularly the emergence of epidemics during winter and spring, pertain exclusively to influenza A(H1N1)pdm09 in Shanghai. Nonetheless, a virus with higher transmissibility could potentially propagate throughout the summer months. In recent years, influenza A(H3N2) has been responsible for several semi-annual outbreaks in Shanghai. [28]. This highlights the complexity of seasonal influenza activity, particularly where co-circulation of various influenza viruses is concerned.

In our modeling analysis, we examined a direct relationship between seasonality and contact patterns without any additional factors, serving as a baseline approximation. For example, the risk of SARS-CoV-2 transmission differs between indoor and outdoor contacts [32] ], which may also apply to influenza. However, our analysis did not distinguish between indoor or outdoor contacts due to the absence of such information in our contact survey.

Furthermore, we observed that seasonality in contact patterns led to seasonal influenza epidemics, but other factors may contribute significantly as well. Previous research has shown that temperature/humidity can affect the survival rate and transmissibility of respiratory viruses [9, 33]. While these factors may independently induce seasonality and can be included separately in a modeling study, we show that seasonality in contact patterns could effectively capture the overall seasonal trends of influenza. Nevertheless, additional studies and analyses able to fully disentangle the intricate relationship between weather and climatic conditions, human contact patterns, and the biological characteristics of pathogens are needed to clarify the inconsistent findings from previous studies that explored the link between climatic conditions and influenza dynamics [8-13].

This study has several limitations concerning the contact survey and the modeling approach. The data on human contact patterns were collected from a single location, so generalizing these findings to areas beyond Shanghai should be approached with caution. However, this is among the few studies exploring the connection between seasonality and human contact patterns, and it can serve as a foundation for guiding future research in this area. Another limitation consists in that we derived conclusions for an entire year despite having only six months of contact data. We addressed this by employing a regression model to predict estimates using temperatures beyond the range of available data. Nevertheless, validating these findings with new data spanning a full year would be highly beneficial, including a deeper investigation of the effect of vacations. The survey was conducted during the pre-COVID era, in the absence of mitigation interventions. Consequentially, it is unknown whether these findings are upheld during or after the COVID-19 pandemic. This analysis considered a constant reporting rate for the influenza season; as observed in other countries [34], it is possible that the reporting rate changes during holiday periods. This could explain the two datapoints outside the 95%CI of model output during the New Year holiday period (Fig. 2A). Finally, regarding the transmission model, the current study uses a simple, homogenous compartmental model to assess patterns of an epidemic consistent with influenza. We did calibrate our model on the 2017-2018 influenza season with consideration for school calendars; however, we did not consider additional factors such as vaccination, age-specific transmission risks, susceptibility to infection, or employment status. Although these factors have the potential to alter transmission dynamics and age-specific outcomes, we do not expect them to determine how contacts change over time and thus should have little impact on the main conclusions of this study.

Our findings can potentially shed light on the seasonality of respiratory illnesses like influenza. Our modeling analysis demonstrates the interaction between the seasonal fluctuations in contact patterns, the introduction of infections, and the subsequent epidemic dynamics. Although this research primarily centers on influenza, the insights gained could be applicable to other respiratory pathogens, such as SARS-CoV-2 and RSV. Nonpharmacologic interventions aimed at limiting the number of contacts were able to reduce SARS-CoV-2 transmission [35-38], showing the effect of changes in contact patterns on epidemic dynamics. However, despite the implementation of interventions contributed to mask seasonal trends in COVID-19, higher rates of SARS-CoV-2 infection and mortality are, were, recorded during the colder months of 2021 [23, 39, 40]. The mechanism proposed in this study could improve our understanding of the transmission patterns of respiratory pathogens and help elucidate their seasonal trends.

## Methods

### Data

#### Contact Survey

The current study is based on data collected from 965 individuals in Shanghai, China who participated in a diary-based contact survey conducted from December 24, 2017, to May 30, 2018. Individual demographic and socioeconomic information of the study participants were collected along with the number of persons with whom they had contact during a 24-hour period before the interview, the date when contacts occurred, and details of each contact (i.e., relationship, location, duration, and type). A full description of the contact survey can be found in *Zhang et al*. [25] and contact diaries are openly available on Zenodo [41].

#### Meteorological data

Meteorological data from October 1, 2017 to September 30, 2018 for Shanghai was obtained from the Hongqiao International Airport Station using an online historical archive of weather reports [42]. Maximum daily temperatures for the study period were matched to the dates when participants completed their contact diaries.

#### Influenza surveillance

We obtained weekly reports of ILIs and laboratory-confirmed influenza infections, as well as total specimens tested, from Shanghai CDC for the 2017-2018 influenza season. In that season, there were 30 sentinel hospitals in Shanghai, including 19 national sentinel hospitals and 11 municipal sentinel hospitals. In each national sentinel hospital, nasopharyngeal swabs were collected from ILI^+^ patients (defined as temperature ≥ 38°C with either cough or sore throat, in the absence of an alternative diagnosis) and placed in sterile viral transport medium for influenza virus testing, resulting in about 20 specimens per hospital per surveillance week. Samples were sent to the Shanghai CDC laboratory for identification of types/subtypes of influenza virus, according to a standard protocol [43]. The weekly number of ILI^+^ cases was multiplied by the weekly proportion of specimens that tested positive for influenza A(H1N1)pdm09 to obtain a better proxy of influenza activity. Similar to previous studies, we refer to this indicator as ILI^+^ [44].

### Statistical Analysis

#### Covariates

Several covariates were included in the analysis to adjust for characteristics and sources of potential influence on human contact patterns. These covariates are age, gender, household size, occupation type, year s lived in Shanghai, weekday, and type of day the diary was completed. Interview responses indicating a participant did not know or was unwilling to answer were recoded as missing.

Age groups were created for the descriptive analysis, stratifying participants into three age groups (0-18, 19-59, and 60+ years old). Age groups were chosen based on dominant social environments (i.e., school, work, and retirement). Gender categories included Male or Female. Occupation type was separated into three categories: Student, Employed, and Not Employed. Participants attending preschool were included in the Student category and the retired participants in the Not Employed category. For the regression analysis, age and household size were normalized by calculating their overall means for each category of the occupation type. These means were then subtracted from the age or household size of individuals in that category to determine their normalized age and their normalized household size.

The variable “years live in Shanghai” was defined as the number of years a participant had been a resident in Shanghai; categories included: < 6 years, 6-10 years, or >10 years/entire life. The weekday variable included three levels: Monday-Friday, Saturday, and Sunday. Type of day was defined as whether the diary was completed on a regular day, irregular day, or New Year holiday. Regular days were considered days of diary completion that did not deviate from the participant’s normal schedule. The irregular day referred to days when a person had significant variations to their normal day schedules (e.g., day off or school holiday). New Year holiday types of the day included participants who reported completing the diaries on irregular days during the New Year holiday, from January 26 to February 22, 2018.

We also included two meteorological covariates: the seasonal trend and the daily temperature variation. We estimated the seasonal trend by fitting a cubic smoothing spline to the maximum daily temperatures between October 1, 2017, and September 30, 2018 (Fig. 1*B*). This accounts for seasonal variations in human behavior throughout the year. Daily temperature variation was defined as the difference between the maximum daily temperatures and the seasonal trend, accounting for the influences of variations of daily temperatures on human behavior.

#### Regression analysis

The effect of seasonal trend and daily temperature variation on total contacts was analyzed using negative binomial regression while adjusting for the following covariates: normalized age, normalized household size, occupation type, gender, weekdays, years lived in Shanghai, and type of day. Observations with a Cook’s Distance greater than 20 times the mean value were considered outliers and excluded from the analysis. Diagnostics were performed to assess regression assumptions. Incidence rate ratios were calculated by exponentiating the coefficients and confidence interval from the regression results (See Supplementary Material for details).

Two sensitivity analyses were performed using alternative meteorological data in Shanghai. All methods remained the same except seasonal trends and daily variations were calculated using average daily temperatures or the daily maximum absolute humidity. We performed a sensitivity analysis where we assessed the effects of seasonal trends and daily temperature variation on community contacts while adjusting for the same covariates (See Supplementary Material).

#### Estimated Number of Contacts

To estimate the daily number of contacts during the study period, we used the temperature data for Shanghai between December 24, 2017, and May 30, 2018 (158 days). The temperature seasonal trend and daily variation were assigned to their true values according to the observed seasonal trend and daily temperature variation for each day. Saturdays and Sundays were identified for each date as well as the New Year holiday period (by using the dummy variables in the regression) according to the calendar. The holiday period was extended to begin on January 22, 2018, to account for the median number of college students whose holiday break began before Shanghai’s public school system. The other regression variables were fixed to specified values for the predictions. In particular, the predictions of the contacts are made for a reference person who is male, employed, has been living in Shanghai for at least 10 years or his entire life, has an average age for an employed person (40 years), and lives in a household of an average size (2 members). We, then, used the regression coefficients to predict the number of contacts in each day. To account for the uncertainty in the coefficients for both, seasonal trend, and daily variation, we performed bootstrap sampling (1,000 draws with replacement) of the coefficients for both variables simultaneously to create simultaneous prediction intervals. Each coefficient draw was used to predict the number of contacts for the defined values of the variables each day. The 95% interquartile range of the resulting vector of predictions were then used as the 95% CI for each day. In addition, we applied these methods to estimate the daily number of contacts outside the time frame of the survey using temperature data from October 1, 2017, to September 30, 2018. We also performed two sensitivity analyses to assess the differences in contact patterns when i) not considering the New Year holiday and ii) considering the summer vacation.

#### Additional Locations

The climate in Shanghai is considered humid, and subtropical; however, the city experiences all four seasons with temperate to cold, damp winters. To evaluate the effect of different climatic conditions on the predicted number of daily contacts, we included two additional locations in China: Beijing and Guangzhou. Beijing’s climate is classified as humid continental where the summers are hot and humid, and the winters are dry and cold, albeit brief. Guangzhou has a humid, subtropical climate, the winters are dry and more temperate. Meteorological data was obtained from the Beijing Capital International Airport Station [45] and the Guangzhou Baiyun International Airport Station [46] from an online historical archive of weather reports. For this sensitivity analysis, we used this data to set the values of the temperature variable and predict the number of daily contacts fixing the regression coeffects at the values estimated for Shanghai. All other covariates were set as for Shanghai. The resulting predicted temporal dynamics of daily contacts was then normalized so that the mean number of contacts over the entire year is the same in all locations. This allows us to compare the variation over time rather than the absolute number of contacts as we are not considering cultural, social, and demographic differences between locations.

### Mathematical Modeling Analysis

#### Infection transmission model

We used the traditional homogenous-mixing SIR model that classified individuals into three compartments: susceptible (*S*), infectious (*I*), and removed (*R*). The following equations were used to simulate the transmission process:

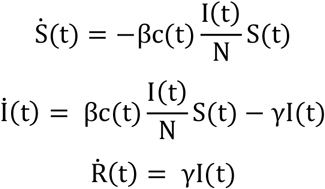

Where N represents the total population of Shanghai (24,860,000 inhabitants as of 2017) [47]. The generation time was set at 3.0 days, in agreement with influenza literature [26]. The contact rate for each day of the study period was derived from the analysis of the contact survey data. The model was also used to estimate the daily net reproduction number by multiplying the contact-dependent basic reproduction number and the fraction of the susceptible population on that day

#### Model Calibration

We defined the likelihood of observing the reported number of ILI^+^ given a binomial distribution with a mean given by the number of weekly infections estimated by the model multiplied by a reporting rate and a given over-dispersion. We then used MCMC to explore this likelihood and estimate the joint posterior distributions of the per-contact transmission risk, the reporting rate, the number of initially infected individuals, and the over-dispersion of the negative binomial distribution used in the likelihood. Metropolis-Hastings sampling was utilized to identify candidate parameters for each iteration of the MCMC. Details for implementing the MCMC process can be found in the Supplementary Material.

#### Reproduction number

The contact-dependent reproduction number was calculated proportional to the number of contacts (regardless of duration, location, and type of contact), using the traditional equation derived for the SIR model [48]:

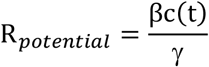

Where:

*c(t)* is the number of contacts at time *t* for a reference individual (see previous section);
β is the per-contact transmission risk as estimated by MCMC;
γ is the recovery rate, which corresponds to the inverse of the generation time for in the traditional SIR model [49].

Statistical analyses were performed in R (version 4.1.0) while the code for the simulation of the mathematical modeling analyses was developed in C

## Data Availability

All data produced in the present study are available upon reasonable request to the authors.

https://doi.org/10.5281/zenodo.3775672

## Ethics statement

Ethics approval was received from the institutional review board of the School of Public Health, Fudan University (IRB no. 2018-01-0659S). Informed consent was obtained from all subjects (from a parent or guardian if the participant was under 18 years of age).

## Acknowledgments

The authors would like to thank Nicole Samay for her assistance in preparing the figures. H.Y. acknowledges funding from the Key Program of the National Natural Science Foundation of China (82130093). A.G.K., P.C.V., M.L., A.V., and M.A. acknowledge support from the Cooperative Agreement no. NU38OT000297 of the Council of State and Territorial Epidemiologists (CSTE). The findings and conclusions in this study are those of the authors and do not necessarily represent the official position of the funding agencies.

## Competing Interest Statement

H.Y. has received research funding from Sanofi Pasteur, GlaxoSmithKline, Yichang HEC Changjiang Pharmaceutical Company, Shanghai Roche Pharmaceutical Company, and SINOVAC Biotech Ltd. M.A. has received research funding from Seqirus. None of this funding is related to this research. The other authors have declared no competing interests.

## Author Contributions

M.L., H.Y., and M.A. designed research. A.G.K., J.Z., and C.J. analyzed the data. A.G.K., J.Z., C.J., M.L., A.V., H.W., H.Y., and M.A. interpreted the results. A.G.K. and M.A. wrote the paper. J.Z., C.J., M.L., P.C.V., M.A.G., A.V., H.W., and H.Y. edited the paper.

## Notes

### Summary of Updates

Revision after receiving reviewers' feedback.

