## Supplementary_Material for "Evaluating seasonal variations in human contact patterns and their impact on the transmission of respiratory infectious diseases"

Allisandra G. Kummer[1,†], Juanjuan Zhang[2,†], Chenyan Jiang[3,†], Maria Litvinova[1], Paulo C. Ventura[1], Marc A. Garcia[4], Alessandro Vespignani[5], Huanyu Wu[3,‡,*], Hongjie Yu[2,‡,*], Marco Ajelli[1,‡,*]

1. Laboratory for Computational Epidemiology and Public Health, Department of Epidemiology and Biostatistics, Indiana University School of Public Health, Bloomington, IN, USA
2. School of Public Health, Fudan University, Key Laboratory of Public Health Safety, Ministry of Education, Shanghai, China
3. Shanghai Municipal Center for Disease Control and Prevention, Shanghai, China
4. Lerner Center for Public Health Promotion, Aging Studies Institute, Department of Sociology, and Maxwell School of Citizenship & Public Affairs, Syracuse University, Syracuse, NY, USA
5. Laboratory for the Modeling of Biological and Socio-technical Systems, Northeastern University, Boston, MA, USA

[†]Co-first author
[‡]Co-senior author
*Corresponding authors

*Seasonal Trend and Contact Patterns* — *3*
**Main Analysis** — **3**
Descriptive Analysis — 3
Correlation Analysis — 3
Incidence Rate Ratios — 5
**Estimated Contacts and Potential Reproduction Numbers** — **5**
Daily Number of Contacts — 5
Additional Analyses: Holidays and Vacations — 6
**Analysis of Community Contacts** — **7**
Descriptive Analysis — 7
Regression Analysis — 7
Incidence Rate Ratio — 8
Estimated Number of Community Contacts — 9
**Sensitivity Analysis: Additional Meteorological Measurements** — **10**
Mean Temperature — 10
Absolute Humidity — 10
Estimated Number of Contacts — 11
**Sensitivity Analysis: Other Locations** — **12**
Estimated Number of Contacts — 12
*Modeling Influenza Spread* — *13*
**Estimated Weekly Number of ILI$^+$ in Shanghai, China** — **13**
**Infection Transmission Model** — **13**
Model Calibration — 13
**Additional Results: Weekly Incidence of New Infections** — **15**

### Seasonal Trend and Contact Patterns

#### Main Analysis

##### Descriptive Analysis

The number of participants per week ranged from 0 to 157, and the total number of contacts each week ranged from 0 to 3,331 contacts (Fig. S1).

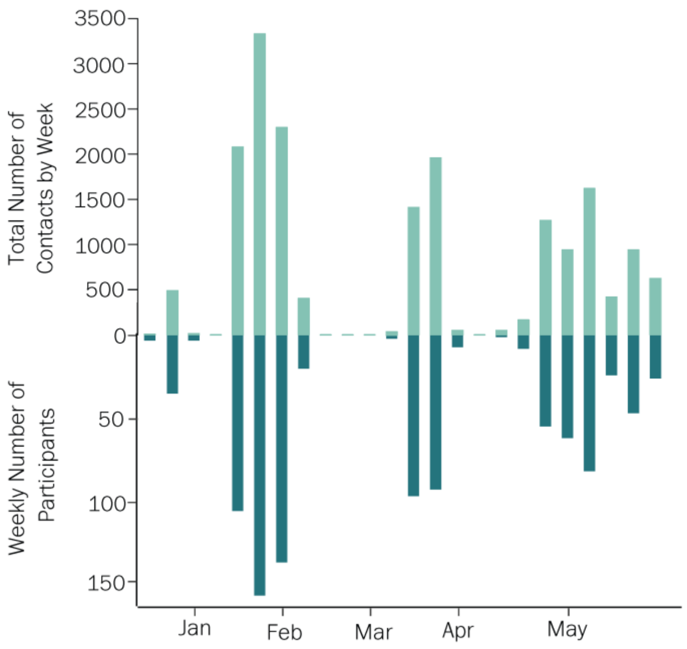


**Figure S1.** Number of total contacts and number of participants interviewed in Shanghai, China by week from December 24, 2017, to May 30, 2018.

Age was defined as the age of the participant on the day they completed the contact diary. Household size was defined as the number of individuals living in the same house as the participant. The average age of participants was 40.7 years old (IQR: 21.0 – 60.0), the average household size was approximately 2 people, not including the participant, and the average maximum temperature in Shanghai during the study period was 15.1 °C (IQR: 7.0 – 25.0) (Tab. S1). For the main analysis, age and household size were normalized by calculating their overall means for each category of the occupation type. These means were then subtracted from the age or household size of participants in that category to determine their normalized age and their normalized household size.

**Table S1.** Descriptive statistics for the variables not reported in the main text.

| | Mean | IQR |
|---|---|---|
| **Age** | 40.7 | (21.0, 60.0) |
| **Household size**[a] | 2.1 | (1.0, 7.0) |
| **Maximum Temperature** | 15.1 | (7.0, 25.0) |

Note. total *N* = 965.
[a]Household size does not include the participant.

##### Correlation Analysis

Collinearities between independent variables were assessed through a Spearman correlation analysis and a Chi-square test (Fig. S2). The seasonal trend and daily temperature variations were inversely connected with the regularity of the daily schedules, reflecting the coinciding of the New Year holidays with the colder season. The rest of the statistically significant correlations were small in size.

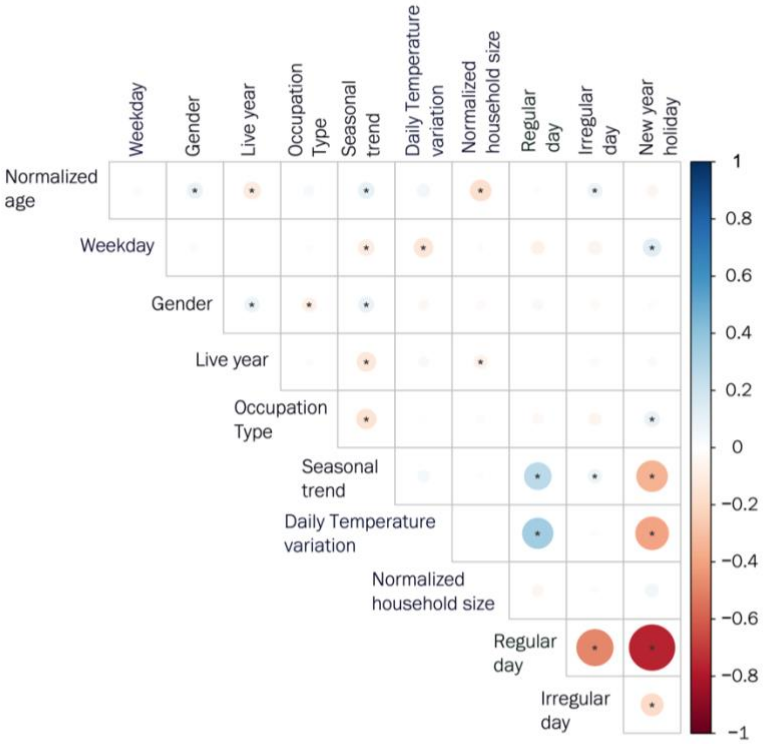


**Figure S2.** Correlation matrix of the independent variables chosen for the main analysis. The * denotes a statistically significant correlation.

#### Other Diagnostics

Observations with Cook's distance greater than 20 times the mean value were considered influential outliers (Fig. S3*A*); therefore, they were excluded from the analysis. Additionally, plotting the fitted and residual values shows a slight pattern, indicating there are unequal variances in the data (Fig. S3*B*).

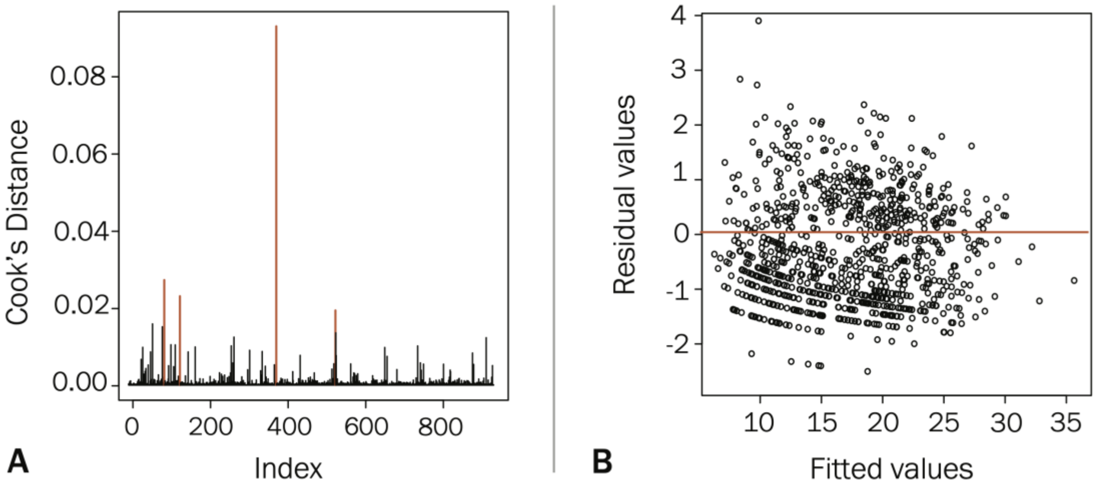


**Figure S3. A.** Cook's distance for each observation was calculated from the regression model for total contacts. Observations in red are considered outliers and were removed for the final regression. **B.** A plot of the residual values vs. the fitted values for the total contacts.

#### Incidence Rate Ratios

Incidence rate ratios (IRR) were calculated by exponentiating the coefficients and confidence interval from the regression results (Fig. S4). Contact rates were lower on Sundays (IRR = 0.817, 95% CI: 0.685 to 0.979) and Saturdays (IRR = 0.674, 95% CI: 0.558 to 0.820) compared to Monday through Friday. Not employed participants had lower rates of contact than those employed (IRR = 0.507, 95% CI: 0.434 to 0.592). As seasonal trends moved from colder to warmer temperatures, the rate of contacts decreased (IRR = 0.987, 95% CI: 0.978 to 0.996). Likewise, increased variations in daily temperatures showed decreased rates in total contacts (IRR = 0.981, 95% CI: 0.966 to 0.996).

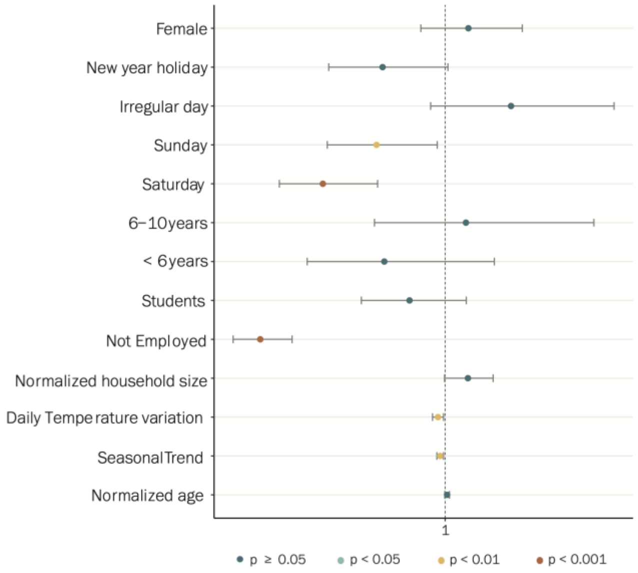


**Figure S4.** Incidence rate ratios of seasonal trend, daily temperature variation, and the covariates of interest for the total contacts.

### Estimated Contacts and Potential Reproduction Numbers

#### Daily Number of Contacts

We estimated the number of contacts for each day from October 1, 2017, to September 30, 2018, using the results of the negative binomial regression. By looking at the daily estimates (instead of grouped by week, as reported in the main text), the effect of weekends on the number of contacts is clearly visible (Fig. S5).

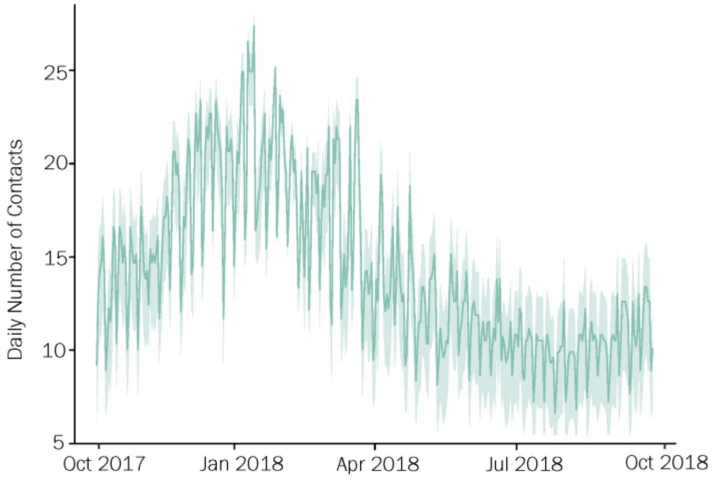


**Figure S5.** Estimated daily number of contacts for each day when seasonal trend, daily variation, and weekday vary while all other variables were fixed between October 1, 2017, and September 30, 2018, without consideration for differences in contact patterns during summer vacation. The line and shaded area represent the mean and 95% CI of the daily values, respectively.

#### Additional Analyses: Holidays and Vacations

We performed two additional analyses for estimating the number of contacts: i) without considering the New Year holiday period and ii) considering the school summer vacation. Without consideration for the New Year holiday (Fig. 6*A*), we see a higher number of contacts during the holiday break period compared to the main analysis with a later peak at the end of January (24.18, 95%CI 19.19-29.60). When considering the summer vacation (Fig. S6*B*), the number of contacts is lower than in the main analysis during the summer vacation period, reaching 7.3 (95%CI: 4.5-9.1) in August.

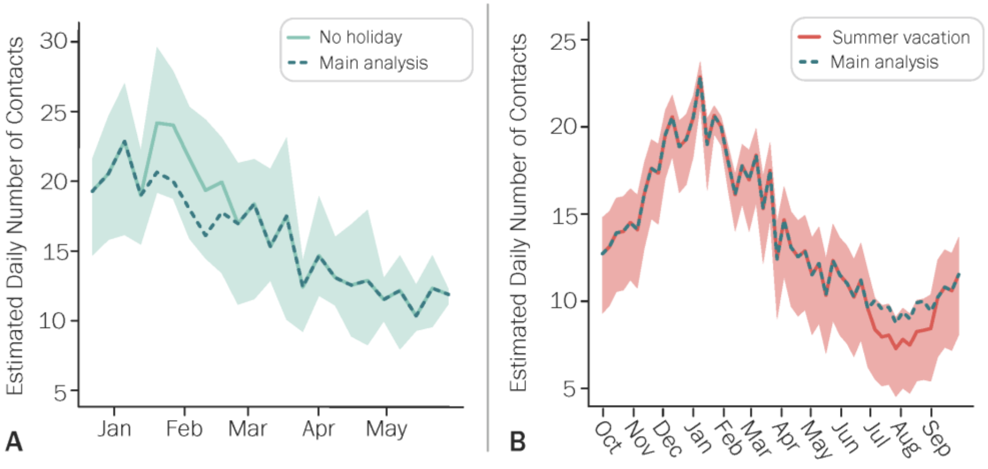


**Figure S6. A.** Estimated daily number of contacts for each week when seasonal trend, daily variation, and weekday vary while all other variables were fixed between December 24, 2017, to May 30, 2018. **B.** Estimated daily number of contacts for each week when seasonal trend, daily variation, and weekday vary while all other variables were fixed from October 1, 2017, to September 30, 2018. The line and shaded area represent the mean and 95% CI of the mean daily values for each season, respectively.

#### Analysis of Community Contacts

##### Descriptive Analysis

Descriptive statistics for the covariates and community contacts are shown in Table S2. Adults 60 years and older reported 8.09 community contacts on average (IQR: 1.0 to 9.0) whereas those 19-59 years had 5.02 community contacts (IQR: 0.0 to 3.0) and participants 0-18 years old had an average of 5.23 community contacts (IQR: 0.0 to 4.0). Employed persons reported a number of community contacts similar to that of students (4.69 vs. 5.13 on average). Participants completing the diaries on Sunday had the most community contacts on average ($\mu$ = 11.7, IQR: 0.0 – 14.0) followed by Saturday with 8.65 community contacts (IQR: 0.0 – 9.0) compared to the working days. Most contacts were had by persons completing the diary on an irregular day ($\mu$ = 8.54, IQR: 0.0 – 11.0) compared to all other types of days.

**Table S2.** Descriptive Statistics for the community contacts by participants' gender, age, occupation, length of time living in Shanghai, and type of day and weekday of participation.

| | Community Contacts | | |
|---|---|---|---|
| | N (%) | Mean | IQR |
| **Total** | 965 (100.0) | 5.9 | (0.0, 5.0) |
| **Gender** | | | |
| Female | 491 (50.9) | 6.5 | (0.0, 6.0) |
| Male | 474 (49.1) | 5.3 | (0.0, 5.0) |
| **Age group** | | | |
| 0-18 | 221 (22.9) | 5.2 | (0.0, 3.0) |
| 19-59 | 477 (49.4) | 5.0 | (0.0, 4.0) |
| 60+ | 267 (27.7) | 8.1 | (1.0, 9.0) |
| **Occupation Type** | | | |
| Students | 252 (26.2) | 5.1 | (0.0, 3.0) |
| Employed | 400 (41.5) | 4.7 | (0.0, 3.0) |
| Not Employed | 307 (31.8) | 8.3 | (1.0, 9.0) |
| Missing | 6 (0.6) | 1.5 | (1.0, 2.0) |
| **Years lived in Shanghai** | | | |
| < 6 years | 47 (4.9) | 6.4 | (0.0, 5.0) |
| 6-10 years | 52 (5.4) | 5.4 | (0.0, 4.0) |
| >10 years/entire life | 862 (89.3) | 5.9 | (0.0, 5.0) |
| Missing | 4 (0.4) | 1.8 | (0.8, 2.0) |
| **Type of Day** | | | |
| Regular day | 812 (84.1) | 5.1 | (0.0, 4.0) |
| New year holiday | 211(21.9) | 7.0 | (0.0, 6.5) |
| Irregular day | 126 (13.1) | 8.5 | (0.0, 11.0) |
| Missing | 27 (2.8) | 7.0 | (3.0, 8.0) |
| **Weekday** | | | |
| Monday - Friday | 705 (73.1) | 4.3 | (0.0, 4.0) |
| Sunday | 143 (14.8) | 11.7 | (0.0, 14.0) |
| Saturday | 117 (12.1) | 8.7 | (0.0, 9.0) |

Note. IQR = Interquartile range.

##### Regression Analysis

Normalized age, not-employed persons, Saturdays, Sundays, irregular days, and new year holidays showed significant increases in expected community contacts compared to Employed individuals, working days, and regular days (Tab. S3). Seasonality showed significant decreases in the number of community contacts. Specifically, for each degree increase in the seasonal trend, the difference in the logs of expected community contacts decreases by 0.025 ($p$ = 0.003, 95% Confidence Interval (CI): -0.048 to -0.013).

**Table S3.** Negative binomial regression model of the effects of seasonal trend and daily temperature variation on community contacts adjusting for the covariates of interest.

| Effect | Community Contacts | | |
|---|---|---|---|
| | Estimate | Pr(>\|z\|) | 95% CI |
| **Seasonal trend** | -0.025 | 0.003** | (-0.048, 0.013) |
| **Daily Temperature variation** | -0.017 | 0.235 | (0.006, 0.030) |
| **Normalized age** | 0.018 | 0.004** | (-0.043, -0.008) |
| **Normalized household size** | 0.012 | 0.843 | (-0.101, 0.127) |
| **Occupation Type** | | | |
| Employed | ref | | |
| Not Employed | 0.921 | 0.0001*** | (0.612, 1.232) |
| Students | -0.093 | 0.541 | (-0.424, 0.243) |
| **Years lived in Shanghai** | | | |
| >10 years/entire life | ref | | |
| < 6 years | 0.095 | 0.734 | (-0.439, 0.695) |
| 6-10 years | -0.161 | 0.547 | (-0.666, 0.404) |
| **Weekday** | | | |
| Monday – Friday | ref | | |
| Saturday | 1.083 | 0.0001*** | (0.720, 1.470) |
| Sunday | 1.611 | 0.0001*** | (1.257, 1.983) |
| **Type of Day** | | | |
| Regular day | ref | | |
| Irregular day | 0.482 | 0.016* | (0.104, 0.891) |
| New year holiday | 0.505 | 0.006** | (0.139, 0.875) |
| **Gender** | | | |
| Male | ref | | |
| Female | 0.162 | 0.187 | (-0.085, 0.408) |

Note. total *N* = 965. *df* = 910. CI =confidence intervals.
*p < 0.05. ** p < 0.01. ***p < 0.001.

#### Incidence Rate Ratio

Incidence rate ratios (IRR) were calculated by exponentiating the coefficients and confidence interval from the regression results (Fig. S7). Community contact rates were higher on Sundays (IRR = 5.006, 95% CI: 3.515 to 7.263) and Saturdays (IRR = 2.953, 95% CI: 2.055 to 4.350) compared to Monday through Friday. Not-employed participants had higher rates of community contacts than those who were employed (IRR = 2.512, 95% CI: 1.844 to 3.428). As seasonal trends moved from colder to warmer temperatures, the rate of community contacts decreased (IRR = 0.975, 95% CI: 0.958 to 0.992).

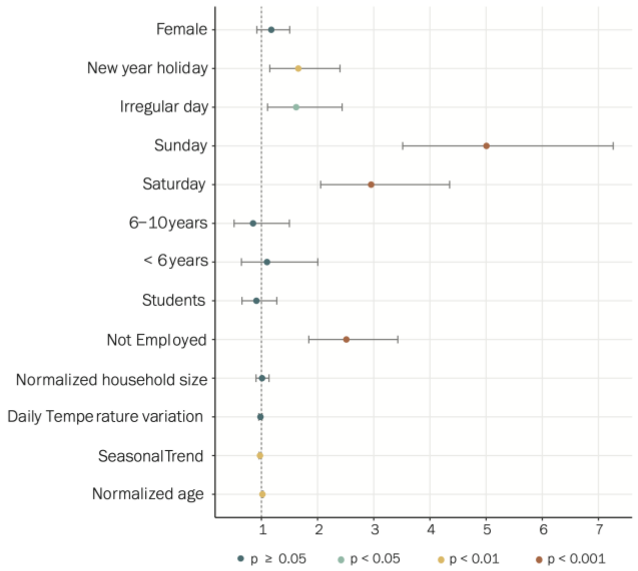

**Figure S7.** Incidence rate ratios of seasonal trend, daily temperature variation, and the covariates of interest for the community contacts.

#### Estimated Number of Community Contacts

There is a slightly decreasing trend in the number of community contacts as daily maximum temperature increases which is heavily influenced by the weekend days (Fig. S8). This indicates both a movement from activities in proximity with others (i.e., indoor aggregations) to activities with lower contacts (i.e., outdoor activities) as well as a distinct difference in weekday vs. weekend contacts.

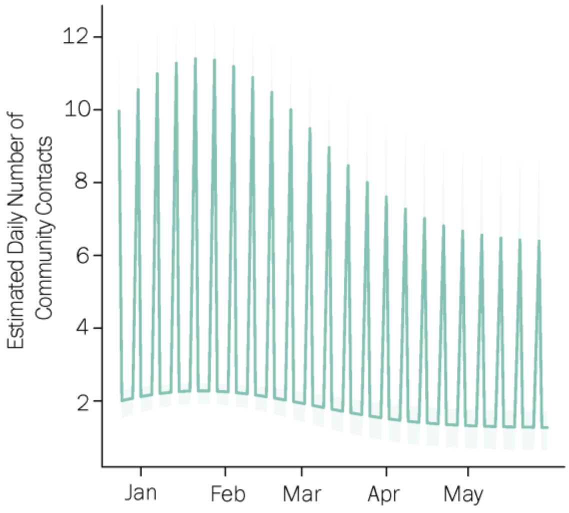


**Figure S8.** Estimated daily number of community contacts accounting for variations in the seasonal trend and weekdays.

#### Sensitivity Analysis: Additional Meteorological Measurements

##### Mean Temperature

In the main analysis, we considered maximum daily temperature as most contacts take place during the daytime, when the temperature is highest. Here we repeat the analysis by considering mean daily temperatures. When the seasonal trend is calculated with respect for mean temperature, not-employed persons, Saturdays, Sundays, and new year holidays showed significant decreases in expected contacts compared to employed individuals, Mondays-Fridays, and regular days (Tab. S4). Seasonality showed significant decreases in the number of total contacts. Specifically, for each degree increase in the seasonal trend, the difference in the logs of expected contacts decreases by 0.016 (p = 0.002, 95% Confidence Interval (CI): -0.026 to -0.006). Likewise, expected total contacts decreases by 0.025 for each degree of daily temperature variation (p = 0.011, 95% CI: -0.045 to -0.005). These results confirm those obtained for the main analysis.

**Table S4.** Negative binomial regression model of the effects of the seasonal trend and daily temperature variation using the average daily temperature on total contacts adjusting for the covariates of interest.

| Effect | **Total Contacts** | | |
|---|---|---|---|
| | Estimate | Pr(>\|z\|) | 95% CI |
| **Seasonal trend** | -0.016 | 0.002** | (-0.026, -0.006) |
| **Daily Temperature variation** | -0.025 | 0.011* | (-0.045, -0.005) |
| **Normalized age** | 0.005 | 0.106 | (-0.001, 0.012) |
| **Normalized household size** | 0.058 | 0.057 | (-0.002, 0.120) |
| **Occupation Type** | | | |
| Employed | ref | | |
| Not Employed | -0.677 | 0.000*** | (-0.832, -0.522) |
| Students | -0.102 | 0.191 | (-0.255, 0.053) |
| **Years lived in Shanghai** | | | |
| >10 years/entire life | ref | | |
| < 6 years | -0.169 | 0.245 | (-0.451, 0.131) |
| 6-10 years | 0.060 | 0.663 | (-0.204, 0.341) |
| **Weekday** | | | |
| Monday – Friday | ref | | |
| Saturday | -0.375 | 0.000*** | (-0.565, -0.178) |
| Sunday | -0.192 | 0.033* | (-0.369, -0.010) |
| **Type of Day** | | | |
| Regular day | ref | | |
| Irregular day | 0.165 | 0.115 | (-0.037, 0.375) |
| New year holiday | -0.210 | 0.039* | (-0.411, -0.008) |
| **Gender** | | | |
| Male | ref | | |
| Female | 0.062 | 0.334 | (-0.066, 0.189) |

Note. total $N$ = 965. $df$ = 910. CI =confidence intervals.
*p < 0.05. ** p < 0.01. ***p < 0.001.

##### Absolute Humidity

When seasonal trend is calculated for absolute humidity, not-employed persons, Saturdays, Sundays, and new year holidays showed significant decreases in expected contacts compared to employed individuals, Mondays-Fridays, and regular days (Tab. S5). Seasonality showed significant decreases in the number of total contacts. Specifically, for each degree increase in the seasonal trend, the difference in the logs of expected contacts decreases by 0.014 (p = 0.010, 95% Confidence Interval (CI): -0.024 to -0.003). Likewise, expected total contacts decreases by 0.033 for each degree of daily temperature variation (p = 0.000, 95% CI: -0.049 to -0.016). These results confirm those obtained for the main analysis.

**Table S5.** Negative binomial regression model of the effects of seasonal trend and daily temperature variation calculated using absolute humidity on total contacts adjusting for the covariates of interest.

| Effect | Total Contacts | | |
|---|---|---|---|
| | Estimate | Pr(>\|z\|) | 95% CI |
| **Seasonal trend** | -0.014 | 0.010* | (-0.024, -0.003) |
| **Daily Temperature variation** | -0.033 | 0.000*** | (-0.049, -0.016) |
| **Normalized age** | 0.006 | 0.088 | (-0.001, 0.012) |
| **Normalized household size** | 0.053 | 0.081 | (-0.007, 0.115) |
| **Occupation Type** | | | |
| Employed | ref | | |
| Not Employed | -0.665 | 0.000*** | (-0.818, -0.511) |
| Students | -0.091 | 0.237 | (-0.244, 0.063) |
| **Years lived in Shanghai** | | | |
| >10 years/entire life | ref | | |
| < 6 years | -0.167 | 0.249 | (-0.449, 0.132) |
| 6-10 years | 0.075 | 0.586 | (-0.188, 0.354) |
| **Weekday** | | | |
| Monday – Friday | ref | | |
| Saturday | -0.387 | 0.000*** | (-0.575, -0.191) |
| Sunday | -0.168 | 0.062 | (-0.344, 0.014) |
| **Type of Day** | | | |
| Regular day | ref | | |
| Irregular day | 0.152 | 0.145 | (-0.049, 0.361) |
| New year holiday | -0.155 | 0.085 | (-0.332, 0.023) |
| **Gender** | | | |
| Male | ref | | |
| Female | 0.060 | 0.342 | (-0.067, 0.187) |

Note. total *N* = 965. *df* = 910. CI =confidence intervals.
*p < 0.05. ** p < 0.01. ***p < 0.001.

#### Estimated Number of Contacts

There is a clear seasonal trend in the weekly number of contacts for mean daily temperature (Fig. S9*B*) and absolute humidity (Fig. S9*C*), reflecting trends seen when using maximum daily temperature (Fig. S9*A*).

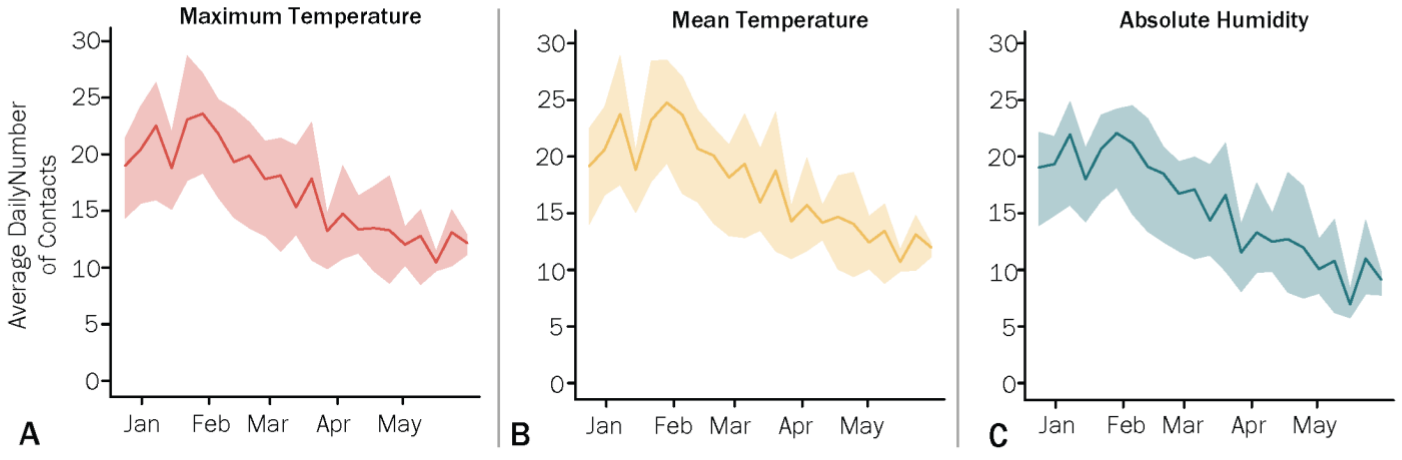


**Figure S9.** Estimated average daily number of contacts for each week when seasonal trend, and daily variation vary with an adjustment for Sundays while all other variables are fixed. The line and shaded area represent the mean and quantiles 0.025 and 0.975 of the daily values. **A.** Maximum temperature. **B.** Mean temperature. **C.** Absolute humidity.

#### Sensitivity Analysis: Other Locations

##### Estimated Number of Contacts

We performed a sensitivity analysis in two additional locations: Beijing and Guangzhou. Seasonal trend and daily variation in temperature were estimated using the maximum daily temperatures in each location from October 1, 2017, to September 31, 2018. The estimated number of contacts for Beijing and Guangzhou show clear seasonal trends that are similar to the trend seen for Shanghai (Fig. S10). However, Beijing demonstrates a greater range of contacts than Shanghai, reaching a maximum of 23.6 contacts (95% CI: 20.9-25.5) in January and a minimum of 8.6 (95% CI: 5.7-11.3) in July (Fig. S10*B*). The estimated number of contacts for Guangzhou is slightly smaller than both locations, reaching a maximum of 21.1 contacts (95% CI: 15.5-25.5) in January and a minimum of 11.6 (95% CI: 7.9-14.7) in August (Fig. S10*C*).

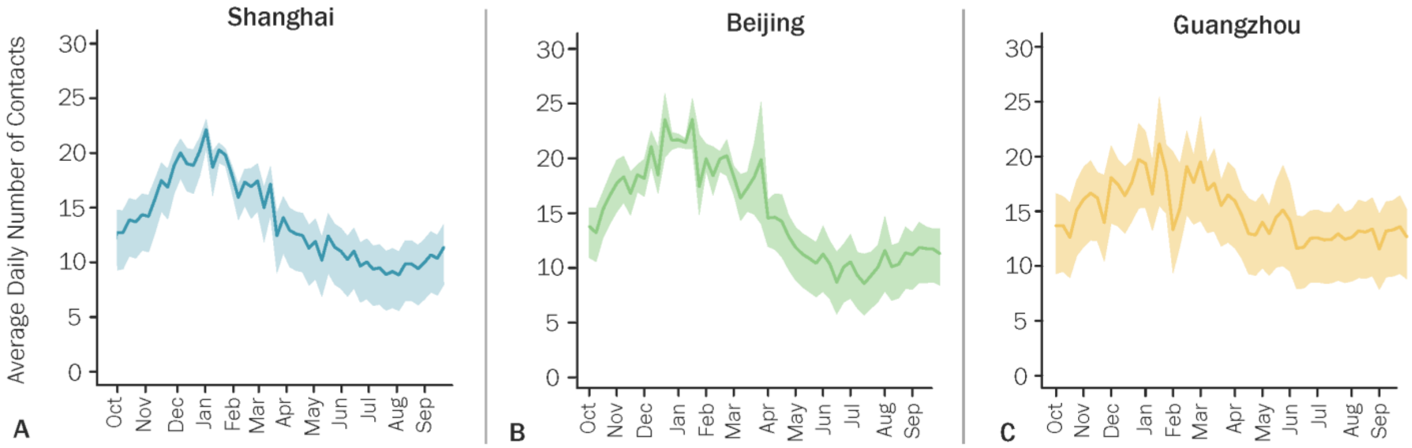


**Figure S10.** Estimated number of contacts when seasonal trend, and daily variation vary with an adjustment for Sundays while all other variables are fixed. The line and shaded area represent the mean and quantiles 0.025 and 0.975 of the daily values. **A.** Shanghai. **B.** Beijing. **C.** Guangzhou.

### Modeling Influenza Spread

#### Weekly Number of ILI+ in Shanghai, China

The following equation was used to approximate the weekly number of ILI+ ($N_{ILI+}$) during the 2017-2018 influenza season.

$$N_{ILI+} = N_{ILI} * \frac{N_{\mathrm{A(H1N1)pdm09}}}{N_{tested}}$$

Where:

- $N_{ILI}$ refers to the weekly number of reported ILI cases;
- $N_{\mathrm{A(H1N1)pdm09}}$ refers to the weekly number of ILI cases that tested positive for A(H1N1)pdm09;
- $N_{tested}$ refers to the weekly number of tested ILI cases.

#### Infection Transmission Model

We first implemented an infection transmission model that incorporates mean total contacts over time ($c_t$) estimated for the number of contacts predicted by the regression model. The following equations were used to simulate the transmission process:

$$\dot{S}_t = -\beta c_t \frac{I_t}{N} S_t$$

$$\dot{I}_t = \beta c_t \frac{I_t}{N} S_t - \gamma I_t$$

$$\dot{R}_t = \gamma I_t$$

$$C_t = \beta c_t \frac{I_t}{N} S_t$$

Where $N$ represents the total population of Shanghai and is set to 24,860,000 which is the population of Shanghai in 2017. The recovery rate corresponds to the inverse of the generation time, which was set to 3.0 days. The contact rate was derived from the analysis of the survey contact data that accounted for the extended holiday season. $C_t$ refers to the cumulative incidence estimated at each time step.

#### Model Calibration

The loglikelihood ($LL$) of observing the number of weekly ILI+ reported in Shanghai during the 2017-2018 influenza season given the weekly number of ILI+ estimated by the infection transmission model was defined as follows:

$$LL = \sum_{t=1}^{18} \log(NB(N_{ILI+}, rK_t, \sigma))$$

Where:

- $N_{ILI+}$refers to the weekly number of ILI+;
- $r$ is the reporting rate;
- $K_t$ is the weekly number of ILI+ estimated by the transmission model;
- $\sigma$ is the over-dispersion of the negative binomial distribution;
- $NB(N_{ILI+}, rK_t, \sigma)$ is the negative binomial probability density function of observing $N_{ILI+}$ from a negative binomial distribution of mean $rK_t$ and over-dispersion $\sigma$.

We used MCMC with Metropolis-Hastings sampling to explore this likelihood and estimate the joint posterior distributions of the parameter vector (transmission risk, number of initial infections imported on December 4, the reporting rate, and the over-dispersion of the negative binomial distribution). We ran chains consisting of 1 million iterations (Fig. S11) and convergence was assessed by selecting multiple initial values of the parameters.

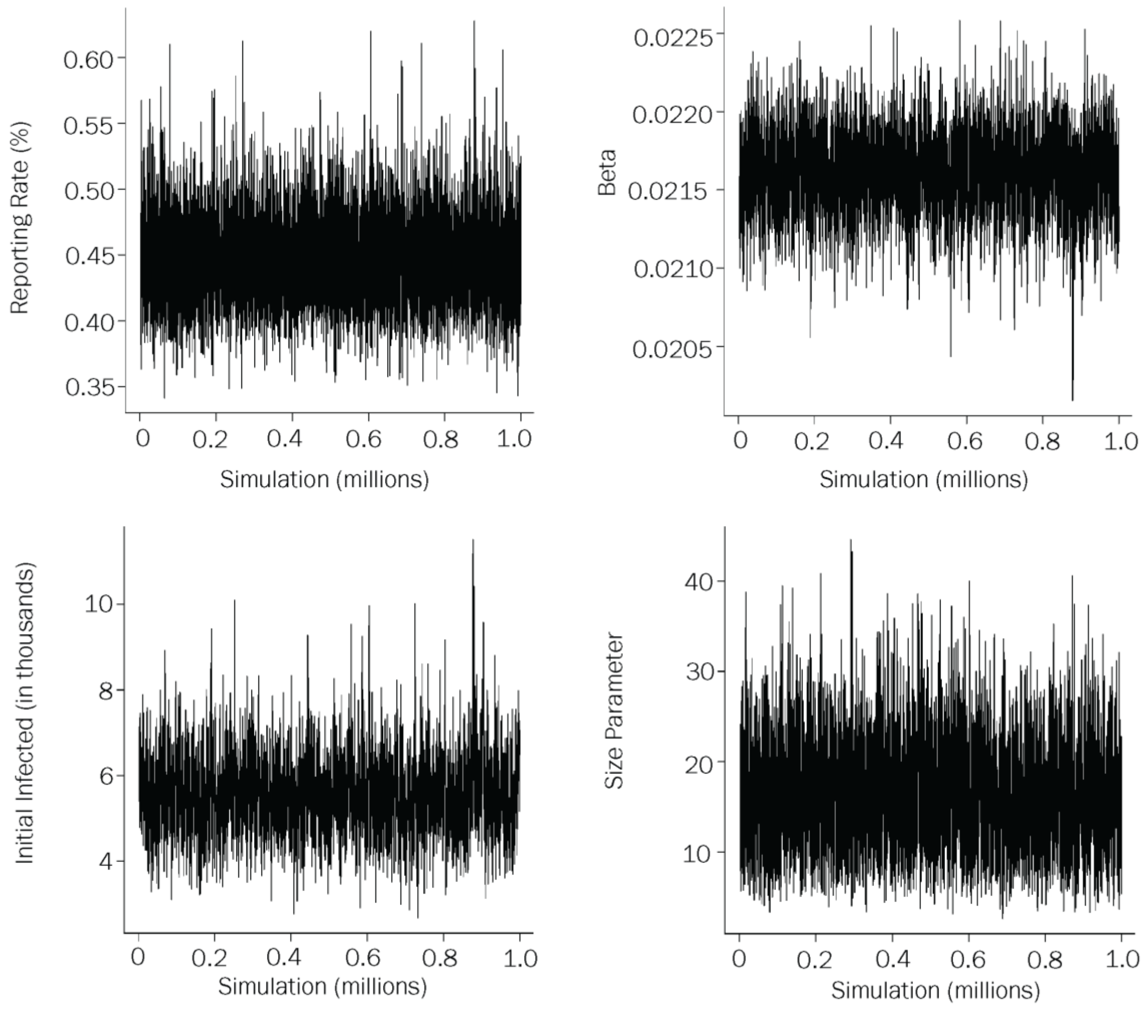


**Figure S11.** Trace plots of the estimated model parameters.

#### Additional Results: Weekly Incidence of New Infections

The results obtained by seeding the epidemic at different times of the year, when the potential reproduction number is above the epidemic threshold, demonstrate seasonal variations in the estimated weekly incidence of new ILI$^+$ infections (Fig. S12). Weekly incidence peaks during the late winter or early spring before declining through the late spring/summer. The weekly incidence for epidemics beginning in February and March remains below 0.25 ILI$^+$ infections during the entire season, while epidemics beginning in November-January see much larger peaks in weekly incidence of ILI$^+$ infections.

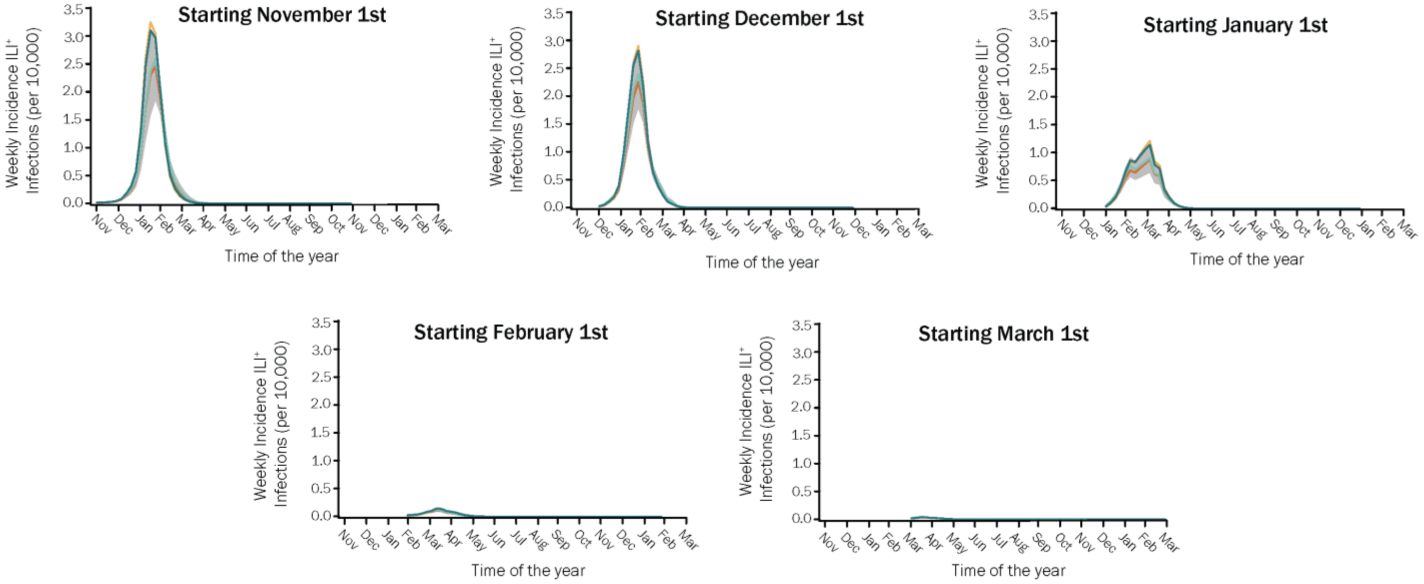


**Figure S12.** Estimated weekly incidence of ILI$^+$ infections in Shanghai, China for epidemics starting at different times of the year, using the estimated posterior distribution of the transmission risk ($\beta$) and initial number of infections for the 2017-2018 influenza season. Lines represent four randomly selected simulations. The shaded area represents the 95% IQR of the distribution.
